# Myocarditis After mRNA-1273 Vaccination: A Population-Based Analysis of 151 Million Vaccine Recipients Worldwide

**DOI:** 10.1101/2021.11.11.21265536

**Authors:** Walter Straus, Veronica Urdaneta, Daina B. Esposito, James A. Mansi, Cesar Sanz Rodriguez, Paul Burton, José M. Vega

## Abstract

**Background:** Growing evidence indicates a causal relationship between SARS-CoV-2 infection and myocarditis. Post-authorization safety data have also identified myocarditis as a rare safety event following mRNA COVID-19 vaccination, most notably among younger adult males and after dose 2. To further evaluate the potential risk after vaccination, we queried the Moderna global safety database to assess the occurrence of myocarditis/myopericarditis among mRNA-1273 vaccine recipients worldwide since first international Emergency Use Authorization issuance.

**Methods:** Reports of myocarditis/myopericarditis entered into the Moderna global safety database from December 18, 2020 to September 30, 2021 were reviewed and classified based on the Brighton Collaboration case definition. The cumulative observed occurrence of myocarditis/myopericarditis was assessed by calculating the reported rate after any known dose of mRNA-1273 according to age and sex. This reporting rate was compared to a population-based incidence rate (US military) to calculate observed-to-expected rate ratios (RR).

**Results:** Through September 30, 2021, a total of 1,439 cases of myocarditis/myopericarditis among approximately 151.1 million mRNA-1273 vaccine recipients were reported to the Moderna global safety database. The overall reporting rate among all vaccine recipients was 0.95 cases per 100,000 vaccine recipients, which was lower than the expected rate from the reference population (2.12 cases per 100,000 vaccine recipients; RR [95% CI]: 0.45 [0.42–0.48]). When stratified by sex and age, observed rates were highest for males aged ≤39 years, particularly those aged 18–24 years (7.40 cases per 100,000 vaccine recipients), which was higher than expected (RR [95% CI]: 3.49 [2.88–4.22]). For males and females aged <18 years, the rate ratio for myocarditis was 1.05 (95% CI, 0.52–2.13) and 0.21 (95% CI, 0.04–0.94), respectively. When considering only cases occurring within 7 days after vaccination, the observed rate was highest for males aged 18–24 years after dose 2 (4.9 cases per 100,000 doses administered).

**Conclusion:** Myocarditis/myopericarditis accounted for 0.4% of adverse events reported to the Moderna global safety database after mRNA-1273 vaccination; rates were higher than expected in males aged 18–24 years, with most occurring by 7 days after dose 2, but were not higher than expected for the overall population of vaccine recipients and were lower than that observed in individuals infected with SARS-CoV-2.

## INTRODUCTION

As the spread of SARS-CoV-2 infection, the causative agent of coronavirus 2019 (COVID-19), continues to pose serious challenges to global health, vaccination in combination with behavioral strategies remain our best approach for controlling the pandemic. In 2020, multiple vaccines against SARS-CoV-2 were developed; most prominent were 2 first-in-class vaccines utilizing messenger RNA (mRNA) technology (mRNA-1273 [SPIKEVAX®; Moderna Inc, Cambridge, MA] and BNT162b2 [COMIRNATY®; BioNTech, Mainz Germany, and Pfizer Inc, New York, NY]). These investigational vaccines were evaluated through numerous clinical studies, including phase 3 studies that randomized approximately 15,000 individuals aged ≥18 years and 22,000 individuals aged ≥16 years to receive mRNA-1273 or BNT162b2, respectively, wherein the vaccines showed no serious safety concerns and were >94% effective against symptomatic infection.^1, 2^

In December 2020, both mRNA-1273 and BNT162b2 received Emergency Use Authorization (EUA) from the US Food and Drug Administration (FDA) for adults ≥18 years and ≥16 years of age, respectively.^3, 4^ Interim recommendations for these age groups were made in December 2020 by the US Centers for Disease Control and Prevention (CDC) Advisory Committee on Immunization Practices (ACIP).^3, 4^ Globally, both mRNA COVID-19 vaccines have since been approved for use in numerous countries, and public health agencies have issued vaccination recommendations for use in adult and adolescent populations.

After a vaccine has been authorized for use, pharmacovigilance surveillance serves as a key source of safety data to provide valuable information for identifying any rare vaccine-related adverse events that may not have been detected in phase 3 studies due to limited sample size. For the mRNA COVID-19 vaccines in particular, the importance of these data is heightened by the fact that there was no prior post-authorization experience for any vaccine developed using the mRNA platform. Since initiation of mRNA COVID-19 vaccination campaigns, a growing body of evidence has largely confirmed the safety findings from clinical studies;^5^ however, reports emerging within 4 to 5 months after EUA also identified cases of vaccinated individuals presenting with myocarditis (with or without pericarditis).^6-9^

The possible link between myocarditis/myopericarditis and mRNA COVID-19 vaccination was first proposed by the Israeli Ministry of Health in April 2021;^10^ subsequently, an Israeli Ministry of Health–appointed epidemiologic team found that between December 2020 and May 2021, 148 cases of predominantly mild myocarditis were reported among approximately 5 million BNT162b2 recipients, primarily in younger males within 30 days after dose 2.^7^ Between January and April 2021, the US military also identified 23 cases of myocarditis among BNT162b2 or mRNA-1273 recipients (after administration of 2.81 million total vaccine doses),^6^ with a higher rate of observed cases versus those expected among military members; similarly to the study in Israel, patients were young males (median age, 25 years) who mostly had symptoms after dose 2.^6^ Of note, myocarditis had not been identified as a safety concern in the phase 3 studies of mRNA-1273 or BNT162b2, including those performed in adolescents.^1, 2, 11, 12^

The initial findings from the Israeli and US military studies prompted researchers, vaccine manufacturers, and public health agencies, including the CDC, to further investigate a potential association between myocarditis and mRNA COVID-19 vaccination.^13-17^ Here, we reviewed the cumulative risk of myocarditis and myopericarditis among global mRNA-1273 vaccine recipients since December 18, 2020 using the Moderna global safety database as the primary data source.

## METHODS

### Data Source and Case Definitions

Worldwide reports of potential myocarditis and myopericarditis entered into the Moderna global safety database during the period of December 18, 2020 (date of first international EUA issuance) to September 30, 2021 were reviewed. The Moderna global safety database is part of an established signal management process that captures spontaneous reports from multiple data sources, including cases reported directly to Moderna, cases reported to regulatory authorities (eg, the US FDA/CDC Vaccine Adverse Event Reporting System [VAERS]), published literature and communications from external sources and (if applicable) business partners, safety data from Moderna-sponsored clinical trials, and other clinical and non-interventional studies. In comparison with clinical trial data, the quality of cases reported to Moderna or through regulatory agencies is more variable and dependent upon details provided by the reporter. This study was based on data collected in the Moderna global safety database, mostly from adverse event reports submitted voluntarily, as part of routine ongoing post-authorization safety surveillance efforts by Moderna, Inc., as required by regulatory authorities; accordingly, a central institutional review board (IRB; Advarra) confirmed this study met criteria for an exemption from IRB oversight under 45 CFR 46.104(d)(4).

The Moderna global safety database was queried for valid case reports of myocarditis and myopericarditis using the Medical Dictionary for Regulatory Activities (MedDRA v24.0) preferred terms shown in **Supplementary Table 1**. Identified cases were classified into 1 of 5 categories, following the Brighton Collaboration myocarditis case definition.^18^ The CDC working case definitions for acute myocarditis^15^ were used for medical review of identified cases. The company causality assessment was provided utilizing the World Health Organization–Uppsala Monitoring Centre standardized case causality assessment.^19^

The global number of mRNA-1273 vaccine recipients was estimated based on information retrieved on October 1, 2021 through the CDC,^20^ the European Centres for Disease Control,^21^ Public Health Agency of Canada,^22^ the Swiss Federal Office of Public Health,^23^ and Our World in Data.^24^ For countries not publishing estimates of mRNA-1273 vaccine use, doses administered were estimated as 50% of doses distributed. United States distributions by age, sex, and doses received supported estimation of stratum-specific vaccine recipient counts. Because BNT162b2 has been authorized for use in US adolescents (aged 12–17 years), it was expected that the large majority of COVID-19 vaccine doses seen in this age group were not mRNA-1273; thus, the total assumed accrued exposure in individuals aged <18 years was limited to 1% of the total.

### Statistical Analysis

Cases of myocarditis and myopericarditis among mRNA-1273 recipients were characterized by age, sex, time to onset after vaccination, and by dose number using descriptive statistics. To calculate the reporting rate of myocarditis, data were stratified and analyzed by age group per available demographic distributions of vaccine recipients published by the US CDC^20^ (<18 years, 18–24 years, 25–39 years, 40–49 years, 50–64 years, 65–74 years, ≥75 years), sex, as well as sex and age group. The reporting rate of myocarditis observed at any time after mRNA-1273 vaccination was calculated as the number of reported myocarditis cases per 100,000 vaccine recipients according to age group and sex. This observed reporting rate of myocarditis was compared with an expected rate from a population-based study of active duty US military.^25^ This reference rate was considered appropriate as a majority of mRNA-1273 doses were distributed in the United States. Although the reference rate was not stratified by age and sex in the published reference, the expected rate of 2.12 cases per 100,000 vaccine recipients was adjusted for lower prevalence among females aged 12–39 years relative to males by factor of 1.73^26^ as presented by the CDC.^27^ The observed reporting rate was divided by the expected rate and presented with an associated 95% confidence interval (CI) calculated as e^(log(IRR)±1.96*SE(log(IRR))))^. Given the observed distribution of latency, dose stratified observed rates of myocarditis occurring within 7 days of an mRNA-1273 vaccination were also estimated as the number of cases per 100,000 doses administered according to age group, sex, and dose; only reported cases with known age, sex, dose number, and time to onset were included in this analysis.

## RESULTS

### Myocarditis and Myopericarditis Cases

From December 18, 2020 to September 30, 2021 a total of 275,252,007 doses of mRNA-1273 were administered globally based on information received from health officials worldwide; during this time period, a total of 332,619 individual case reports (containing 1,284,222 adverse events, of which 178,301 were considered serious) were entered into the Moderna global safety database. Note that a single case can contain more than one reported event.

Of these 332,619 reported individual cases, a total of 1,439 were cases of myocarditis or myopericarditis (0.4% of reported cases). Of the 1,439 reported cases, 1,136 (79%) were reported by a medically qualified healthcare professional involved in the patient’s care; 1,434 (99.6%) cases were considered serious, including 21 (1.5%) that had a fatal outcome (detailed in the Supplement). As of the date of report, there were 692 cases (48.1%) where event status was reported as recovered (314 cases; 21.8%), recovered with sequelae (13 cases; 0.9%), or recovering (365 cases; 25.4%); 378 cases (26.3%) reported an outcome of not recovered and the outcome was not reported in 348 cases (24.2%).

The majority of cases originated from the EEA (584 cases; 40.6%) and the United States (560 cases; 38.9%); 116 cases (8.1%) were reported from Asia, 92 (6.4%) from Switzerland, 78 (5.4%) from the United Kingdom, and <1% were from Canada (8 cases, 0.6%) and the Middle East (1 case; 0.1%). The median age of patients with reported myocarditis or myopericarditis was 27 years (range, 13–94 years). Cases were most frequently reported among males (1,117 cases; 77.6%), with the majority of cases involving males aged 18–39 years (887 cases; 61.6%; **Figure 1**). Among females, cases primarily occurred in those aged 18–29 years (97 cases; 6.7%).

**Figure 1.**
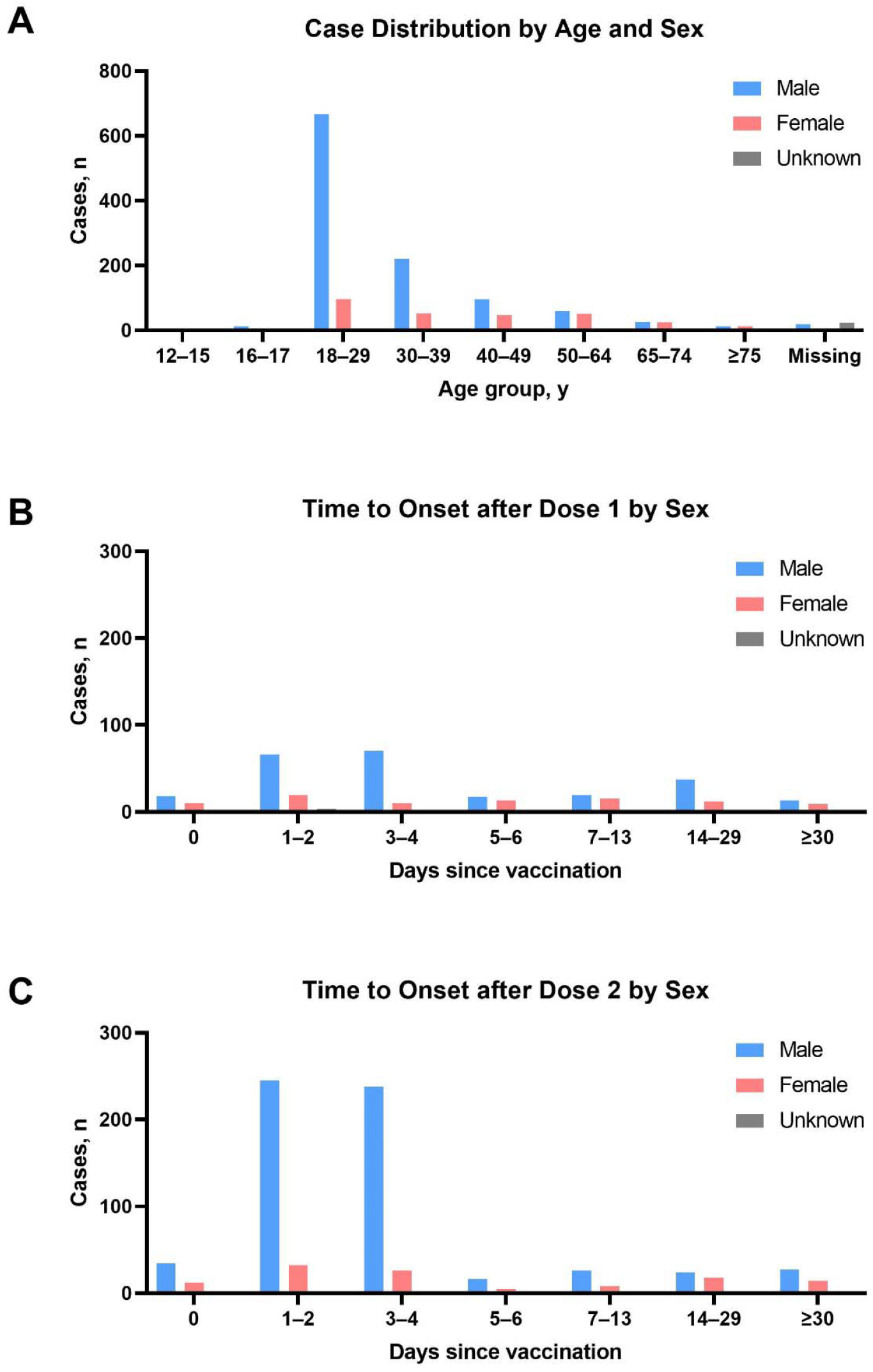
Distribution and time to onset of myocarditis and myopericarditis cases after mRNA-1273 vaccination. (A) Distribution of the 1,439 reported myocarditis and myopericarditis cases among mRNA-1273 vaccine recipients according to age and sex. A total of 47 cases were missing age and/or sex information. (B) Timing of the 331 reported cases of myocarditis and myopericarditis after dose 1 of mRNA-1273. (C) Timing of the 729 reported cases of myocarditis and myopericarditis after dose 2 of mRNA-1273. A total of 376 cases were missing dose number and/or sex information.

Of the 1,439 reported cases of myocarditis and myopericarditis, 729 cases (50.7%) were reported to have occurred after dose 2, 331 cases (23.0%) after dose 1, and 3 cases (0.2%) after dose 3 (**Table 1**). A total of 376 cases (26.1%) had missing dose information. Onset of myocarditis after vaccination was typically within 6 days after any dose (840 cases; 58.4%; **Figure 1**), with 611 cases (42.5%) occurring within 6 days after dose 2.

**Table 1.** Time to Onset of Myocarditis and Myopericarditis Cases by mRNA-1273 Dose Number.

| Time to Onset, days | Myocarditis/Myopericarditis Cases, n (%<sup>a</sup>) |  |  |  |
| --- | --- | --- | --- | --- |
|  | Dose 1 | Dose 2 | Dose 3 | Any Dose |
| <7 | 226 (15.7) | 611 (42.5) | 3 (0.2) | 840 (58.4) |
| 7–29 | 83 (5.7) | 77 (5.4) | 0 | 160 (11.1) |
| ≥30 | 22 (1.5) | 41 (2.8) | 0 | 63 (4.4) |
| **Total<sup>b</sup>** | 331 (23.0) | 729 (50.7) | 3 (0.2) | 1,439 |

<sup>a</sup>Percentage based on the total number of reported myocarditis and myopericarditis cases after any dose of mRNA-1273.
<sup>b</sup>A total of 376 cases (26.1%) contained insufficient information to determine time to onset after mRNA-1273.

Among those with information available on symptom onset, the median time to onset was 3 days; among those with information on symptom duration, the median duration of events was 5 days.

### Observed Versus Expected Rates of Myocarditis and Myopericarditis

The 1,439 cases of myocarditis and myopericarditis reported to the Moderna global safety database between December 18, 2020 and September 30, 2021 corresponded to an overall observed reporting rate of 0.95 cases per 100,000 vaccine recipients. The observed reporting rate of myocarditis among mRNA-1273 recipients varied by age and sex but generally occurred at a rate of <1 case per 100,000 vaccine recipients (**Table 2**). Myocarditis reporting rates were 1.56 cases per 100,000 vaccine recipients for males and 0.37 cases per 100,000 vaccine recipients for females. Across age groups, reporting rates of myocarditis were highest for individuals 18–24 years of age (3.98 cases per 100,000 vaccine recipients). When stratified by age and sex, younger males aged ≤39 years had the highest reported rates of myocarditis, particularly those aged 18–24 years (7.40 cases per 100,000 vaccine recipients); among females, rates were also highest for those aged 18–24 years (0.89 cases per 100,000 vaccine recipients).

**Table 2.** Observed Versus Expected Rates of Myocarditis and Myopericarditis Among mRNA-1273 Recipients (per 100,000 Vaccine Recipients): Moderna Global Safety Database From December 18, 2020, to September 30, 2021.

|  | mRNA-1273<br>Recipients<sup>a</sup> | Observed |  | Expected |  | Rate Ratio<br>(95% CI) |
| --- | --- | --- | --- | --- | --- | --- |
|  |  | Cases (N) | Rate<sup>b</sup> | Cases<sup>c</sup> (N) | Rate<sup>b</sup> |  |
| All recipients |  |  |  |  |  |  |
| All ages | 151,170,178 | 1439 | 0.95 | 3204.8 | 2.12 | 0.45 (0.42 –0.48) |
| <18<sup>d</sup> years | 1,511,702 | 18 | 1.19 | 25.3 | 1.67 | 0.71 (0.39–1.30) |
| 18–24<sup>d</sup> years | 13,605,316 | 541 | 3.98 | 227.6 | 1.67 | 2.38 (2.04–2.78) |
| 25–39<sup>d</sup> years | 33,257,439 | 500 | 1.50 | 556.3 | 1.67 | 0.90 (0.80–1.01) |
| 40–49 years | 22,675,527 | 144 | 0.64 | 480.7 | 2.12 | 0.30 (0.25–0.36) |
| 50–64 years | 39,304,246 | 112 | 0.28 | 833.3 | 2.12 | 0.13 (0.11–0.16) |
| 65–74 years | 22,675,527 | 51 | 0.22 | 480.7 | 2.12 | 0.11 (0.08–0.14) |
| ≥75 years | 15,117,018 | 26 | 0.17 | 320.5 | 2.12 | 0.08 (0.05–0.12) |
| Male recipients |  |  |  |  |  |  |
| All ages | 71,654,664 | 1117 | 1.56 | 1519.1 | 2.12 | 0.74 (0.68–0.79) |
| <18 years | 716,547 | 16 | 2.23 | 15.2 | 2.12 | 1.05 (0.52–2.13) |
| 18–24 years | 6,448,920 | 477 | 7.40 | 136.7 | 2.12 | 3.49 (2.88–4.22) |
| 25–39 years | 15,764,026 | 410 | 2.60 | 334.2 | 2.12 | 1.23 (1.06–1.42) |
| 40–49 years | 10,748,200 | 97 | 0.90 | 227.9 | 2.12 | 0.43 (0.34–0.54) |
| 50–64 years | 18,630,213 | 59 | 0.32 | 395.0 | 2.12 | 0.15 (0.11–0.20) |
| 65–74 years | 10,748,200 | 26 | 0.24 | 227.9 | 2.12 | 0.11 (0.08–0.17) |
| ≥75 years | 7,165,466 | 13 | 0.18 | 151.9 | 2.12 | 0.09 (0.05–0.15) |

| Female recipients |  |  |  |  |  |  |
| --- | --- | --- | --- | --- | --- | --- |
| All ages | 79,515,513 | 292 | 0.37 | 1380.9 | 1.74 | 0.21 (0.19–0.24) |
| <18<sup>d</sup> years | 795,155 | 2 | 0.25 | 9.7 | 1.23 | 0.21 (0.04–0.94) |
| 18–24<sup>d</sup> years | 7,156,396 | 64 | 0.89 | 87.7 | 1.23 | 0.73 (0.53–1.01) |
| 25–39<sup>d</sup> years | 17,493,413 | 86 | 0.49 | 214.4 | 1.23 | 0.40 (0.31–0.52) |
| 40–49 years | 11,927,327 | 46 | 0.39 | 252.9 | 2.12 | 0.18 (0.13–0.25) |
| 50–64 years | 20,674,033 | 51 | 0.25 | 438.3 | 2.12 | 0.12 (0.09–0.16) |
| 65–74 years | 11,927,327 | 25 | 0.21 | 252.9 | 2.12 | 0.10 (0.07–0.15) |
| ≥75 years | 7,951,551 | 13 | 0.16 | 168.6 | 2.12 | 0.08 (0.04–0.14) |

<sup>a</sup>Rates presented per 100,000 vaccine recipients; vaccine recipients extrapolated based on the proportion of vaccine administrations given as dose 1 versus dose 2 in the United States.
<sup>b</sup>Calculated as the rate of myocarditis and myopericarditis cases per 100,000 vaccine recipients.
<sup>c</sup>Number of cases based on data from the US military.<sup>25</sup>
<sup>d</sup>Expected cases among females 12–39 years of age were adjusted for a lower prevalence to males by a factor of 1.73<sup>26</sup> based on presentation from the US CDC.<sup>27</sup>

Comparing observed rates of myocarditis and myopericarditis after mRNA-1273 vaccination to the expected rates based on background incidence in the US military data^25^ resulted in an overall rate ratio of 0.45 (95% CI, 0.42–0.48; **Table 2**). Across age groups, a higher-than-expected rate of myocarditis was observed for those individuals 18–24 years of age (2.38 [95% CI, 2.04–2.78). When examining the data further by sex, a higher-than-expected rate was observed for males 18–24 years of age (3.49 [95% CI, 2.88–4.22]); rate ratios were 1.05 (95% CI, 0.52–2.13) for males <18 years and 1.23 (95% CI, 1.06–1.42) for males 25–39 years of age. Females aged <18 and 18–24 years had rate ratios of 0.21 (95% CI, 0.04–0.94) and 0.73 (95% CI, 0.53–1.01), respectively.

### Myocarditis and Myopericarditis Rates Within 7 Days of mRNA-1273 Vaccination

A total of 840 reported cases of myocarditis and myopericarditis had information available on the timing of myocarditis onset and the dose number of mRNA-1273 received before the occurrence of the event. When analysis of the cases was restricted to onset within 7 days after any vaccination dose, the observed rate of myocarditis per 100,000 doses administered was generally higher for all vaccine recipients after dose 2 **(Table 3**). Overall, the highest rates within 7 days after vaccination were reported after dose 2 in younger males aged ≤39 years, particularly among those aged 18–24 years (4.9 cases per 100,000 doses administered).

**Table 3.** Reported Rate of Myocarditis and Myopericarditis Within 7 Days of mRNA-1273 According to Age and Dose Number (per 100,000 Doses Administered): Moderna Global Safety Database From December 18, 2020, to September 30, 2021.

|  | Reported Rate of Myocarditis/Myopericarditis<br>Within 7 Days<sup>a</sup> |  |
| --- | --- | --- |
|  | Dose 1 | Dose 2 |
| **All recipients** |  |  |
| <18 years | 0.26 | 0.56 |
| 18–24 years | 0.65 | 2.56 |
| 25–39 years | 0.28 | 0.78 |
| 40–49 years | 0.09 | 0.33 |
| 50–64 years | 0.07 | 0.09 |
| 65–74 years | 0.02 | 0.05 |
| ≥75 years | 0.05 | 0.02 |
| **Male recipients** |  |  |
| <18 years | 0.56 | 1.02 |
| 18–24 years | 1.15 | 4.91 |
| 25–39 years | 0.48 | 1.44 |
| 40–49 years | 0.14 | 0.52 |
| 50–64 years | 0.05 | 0.14 |
| 65–74 years | 0.03 | 0.07 |
| ≥75 years | 0.03 | 0.02 |
| **Female recipients** |  |  |
| <18 years | 0 | 0.15 |
| 18–24 years | 0.20 | 0.44 |
| 25–39 years | 0.09 | 0.19 |
| 40–49 years | 0.05 | 0.15 |
| 50–64 years | 0.07 | 0.05 |
| 65–74 years | 0.01 | 0.03 |
| ≥75 years | 0.06 | 0.03 |

## DISCUSSION

To further understand the potential association between myocarditis and mRNA-1273 vaccination, we evaluated the worldwide rates of myocarditis and myopericarditis among vaccine recipients as cumulatively reported to the Moderna global safety database through September 30, 2021. Since the first EUA (December 18, 2020), this database has collected spontaneous adverse events following mRNA-1273 vaccination as reported from multiple data sources. In this analysis, the vast majority of reports were received from regulatory authorities, primarily originating from the United States and EEA. Overall, the rate of myocarditis among vaccine recipients during this time period was low (1,439 cases; 0.95 cases per 100,000 vaccine recipients); however, as with recent reports in the literature,^6, 13, 14, 16, 17, 28^ higher rates were observed for younger males ≤39 years of age, particularly after dose 2. Males aged 18–24 years had the highest rate of myocarditis after vaccination (7.40 cases per 100,000 vaccine recipients), 3.5-fold higher than the expected rate in this age group.

Although myocarditis after vaccination is generally considered a rare safety event, infrequent cases have been previously reported for certain vaccines.^29^ In addition to the recent reports of myocarditis after mRNA-COVID-19 vaccination,^6, 13, 14, 16, 17, 28, 30^ myocarditis has also been observed following smallpox vaccination with a live vaccinia virus vaccine. In 1 study, a rate of 7.8 myocarditis cases per 100,000 smallpox vaccine recipients was reported over a 30-day window, a 3.6-fold higher rate than normally expected.^31^ In the general population, there are numerous infectious and non-infectious causes of myocarditis, with infection by any of several viruses currently representing the most common cause.^32^

Since the emergence of SARS-CoV-2, growing evidence also indicates a causal relationship between natural SARS-CoV-2 infection and myocarditis, with 1 recent analysis from the CDC estimating that the risk of myocarditis among hospitalized persons was 15.7 times higher for patients with COVID-19 than those without COVID-19.^33^ Further, the risk of developing myocarditis from SARS-CoV-2 infection was higher for males than females and highest for children (<16 years of age) and older adults (≥50 years of age).^33^ The prominent role SARS-CoV-2 infection plays in myocarditis for young males was also highlighted in a separate analysis, which estimated the rate of myocarditis from primary infection as 450 per million among males 12–17 years of age, a rate approximately 6 times higher than that observed after mRNA COVID-19 vaccination.^34^

Notably, a population-based cohort study of approximately 2.4 million patients aged ≥18 years observed 15 cases of confirmed myocarditis after any dose of a mRNA COVID-19 vaccine (2 cases after dose 1 and 13 cases after dose 2), for an incidence of 0.08 cases per 100,000 first doses and 0.58 cases per 100,000 second doses; acute myocarditis was described as a rare event.^14^ All reported cases occurred in younger males (median age of 25 years) who were hospitalized and had symptoms resolve with conservative management. The US FDA recently presented an assessment of myocarditis/pericarditis rates using the FDA Biologics and Effectiveness Safety (BEST) active surveillance system, a database consisting of 4 health claims data sources with a total 76.5 to 89.5 million enrollees per year.^35^ Within the first 7 days after administration of any dose of an mRNA COVID-19 vaccine (eg, mRNA-1273 or BNTN162b2), the incidence rate of myocarditis/pericarditis per 1 million person-days was generally low for all age groups; rates were highest for males aged 18–25 years after dose 2. At the October 21, 2021 CDC ACIP meeting, the COVID-19 Vaccine Safety Technical (VaST) Work Group summarized the available data to date on myocarditis rates after mRNA COVID-19 vaccination from multiple worldwide safety monitoring systems,^36^ which indicated that myocarditis was associated with both mRNA-1273 and BNT162b2 vaccination among adolescents and young adults, more frequently among males. The COVID-19 subcommitee of the WHO Global Advisory Committee on Vaccine Safety (GACVS) also recently reviewed available evidence from multiple countries and noted that while some data suggest increased incidence of myocarditis in young males after dose 2 of mRNA-1273 versus BNT162b2, other data do not support this finding, and the overall risk of myocarditis is low.^37^

Several large US surveillance systems have shown comparable risk of myocarditis between mRNA-1273 and BNT162b2 (eg, VAERS, FDA BEST System, and Department of Veterans Affairs active surveillance Rapid Cycle Analysis for COVID-19 vaccines).^36^ For example, the CDC COVID-19 Vaccine Task Force provided the reporting rate of myocarditis after mRNA-1273 and BNT162b2 vaccination among males 18–24 years of age as 3.68 and 3.85 cases per 100,000 second doses administered, respectively, based on data from the US VAERS safety passive monitoring system.^38^ However, a recent analysis from the Vaccine Safety Datalink (VSD) estimated that there was an excess 9.7 cases of myocarditis/myopericarditis per million doses of mRNA-1273 versus BNT162b2 among 18–39-year-olds (adjusted rate ratio [95% CI]: 2.28 [1.00–5.22]; 2-sided p-value: 0.049).^39^ Of note, the VSD analysis was based on a small number of cases within 7 days after dose 2 (mRNA-1273: 14 cases [810,839 total second doses]; BNT162b2: 12 cases [1,256,525 total second doses]).^39^

Most importantly, it is critical to put these rare and generally mild events in the context of the number of cases of myocarditis prevented by the COVID-19 vaccines. Numerous studies have demonstrated the robust effectiveness of mRNA 1273 in real life practice.^40-43^ Further, a report by members of the CDC COVID-19 Response Team and colleagues, summarized at the ACIP meeting on June 23, 2021, estimated that over a 120-day period, 45 to 56 cases of myocarditis per million dose 2 vaccinations were predicted to occur in males aged 18–24 years, whereas mRNA COVID-19 vaccination would simultaneously prevent 12,000 COVID-19 cases, 530 hospitalizations, 127 admissions to the intensive care unit, and 3 deaths.^15^ Based upon extensive review of this and other available evidence, the authors concluded that the benefit of mRNA COVID-19 vaccination clearly outweighed the risk of myocarditis in all recommended age groups, including younger male adolescents at heightened risk for myocarditis after vaccination.^15^ Of note, reported cases of myocarditis and myopericarditis after vaccination are generally mild, self-limiting, and resolved using conservative treatment.^13-15, 17, 37, 44^

The current analysis utilized information submitted to the Moderna global safety database, which continuously receives safety reports from a large, geographically diverse population, enabling detection and assessment of rare safety events on an ongoing and up-to-date basis. A key limitation of this analysis is that the information from the Moderna global safety database is primarily derived from passive, spontaneous adverse event reporting, which lack a denominator (to clearly define the number of individuals who have received the vaccine) and often provide limited details on the clinical features and outcomes of reported myocarditis cases. As such, chart reviews and follow-ups on case resolutions were precluded.

In conclusion, our findings demonstrate that myocarditis in individuals receiving mRNA 1273 is a rare event; previous reports suggest these cases are likely generally mild and self-limiting.^13-15, 17, 44^ As noted in a recent update based on data presented to the ACIP, the benefits of mRNA COVID-19 vaccination (ie, prevention of COVID-19, hospitalization, intensive care unit admissions, and death) clearly outweigh the potential harm of vaccine-related myocarditis.^15^ In safely and effectively preventing COVID-19 disease and its complications (including myocarditis, a natural complication of SARS-CoV-2 infection), mRNA COVID-19 vaccines thus remain essential for controlling the COVID-19 pandemic.

## Data Availability

Patient-level data reported in this study are not shared publicly but they are shared fully with regulatory agencies.

## Funding

This study was funded by Moderna, Inc.

## Conflicts of Interest

All authors are employees of and shareholders in Moderna, Inc.

## Acknowledgments

We thank Margot Stam Moraga, Vasudev Bhupathi, Alicia Pucci, and Melissa Rossi of Moderna, Inc., for their contributions to the extraction, analysis, and interpretation of the data. Medical writing and editorial assistance were provided by MEDiSTRAVA in accordance with Good Publication Practice (GPP3) guidelines, funded by Moderna, Inc., and under the direction of the authors.

## SUPPLEMENT

### Supplementary Results

Of the 21 fatal reports of myocarditis or myopericarditis, 15 were male (71.4%) and 6 were female (28.6%); the median age was 59 years (range, 17–85 years). Five fatal reports occurred after dose 1 with a time to onset (TTO) of 0–17 days; 4 reports were after dose 2 with a TTO of 1–4 days. Twelve fatal reports occurred after an unknown dose number and for which a TTO was also not reported. These reports were heavily confounded by patient medical histories, including rheumatoid arthritis, pericardial drainage, hypothyroidism, Prinzmetal angina, hypertension, coronary artery disease, chronic left ventricular failure, chronic right ventricular failure, diabetes mellitus, prostate cancer, *Clostridium difficile* infection, myocardial infarction, cerebrovascular accident, myocardial fibrosis, cardiogenic shock, and pneumonia pneumococcal, among others.

**Supplementary Table 1.**
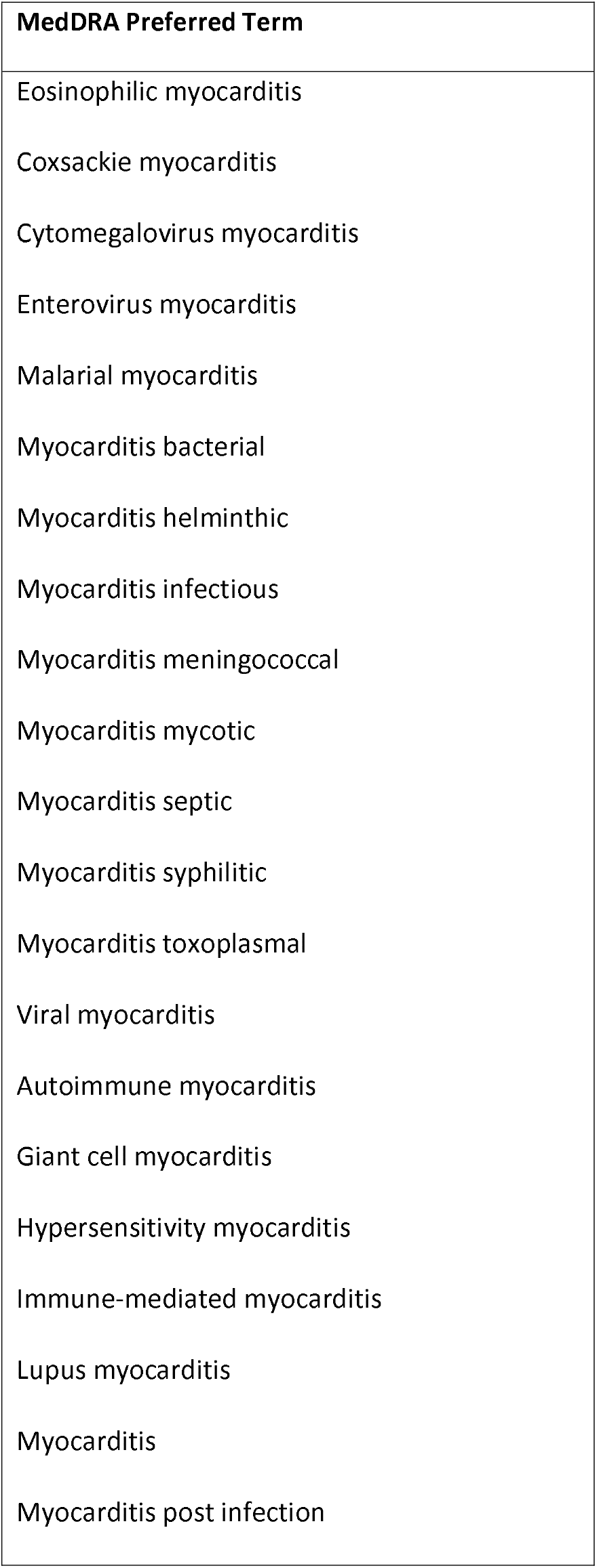
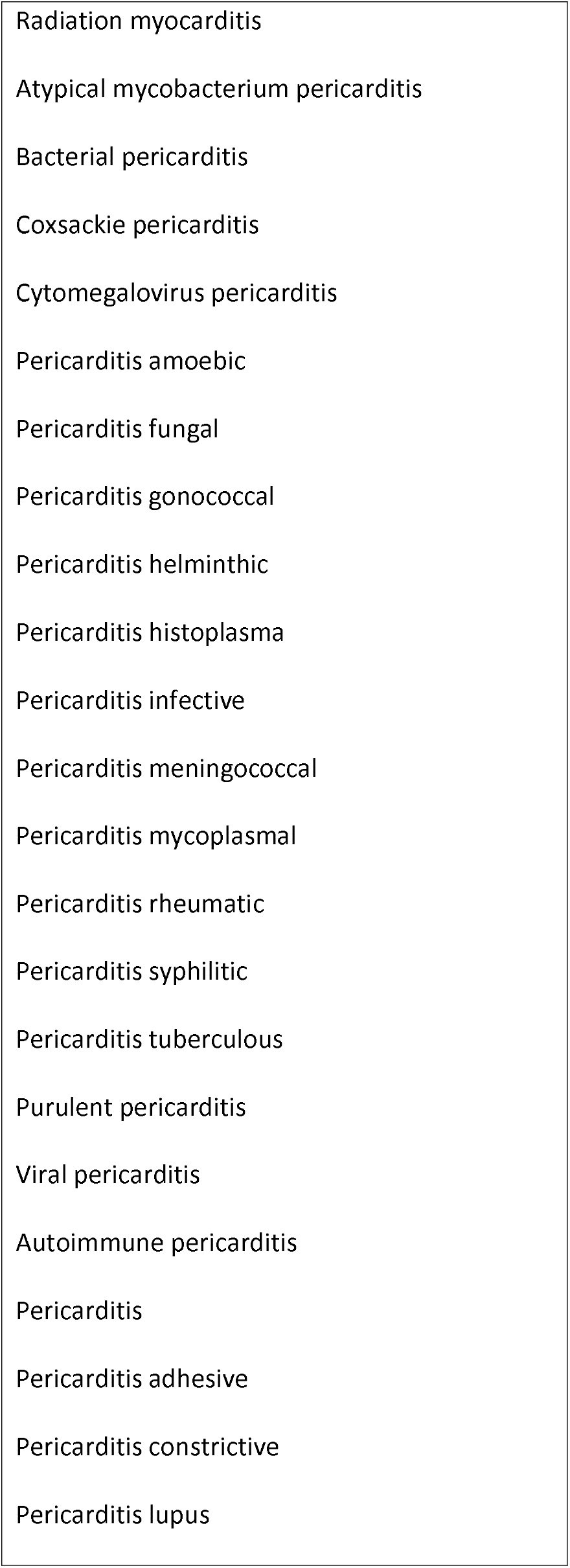
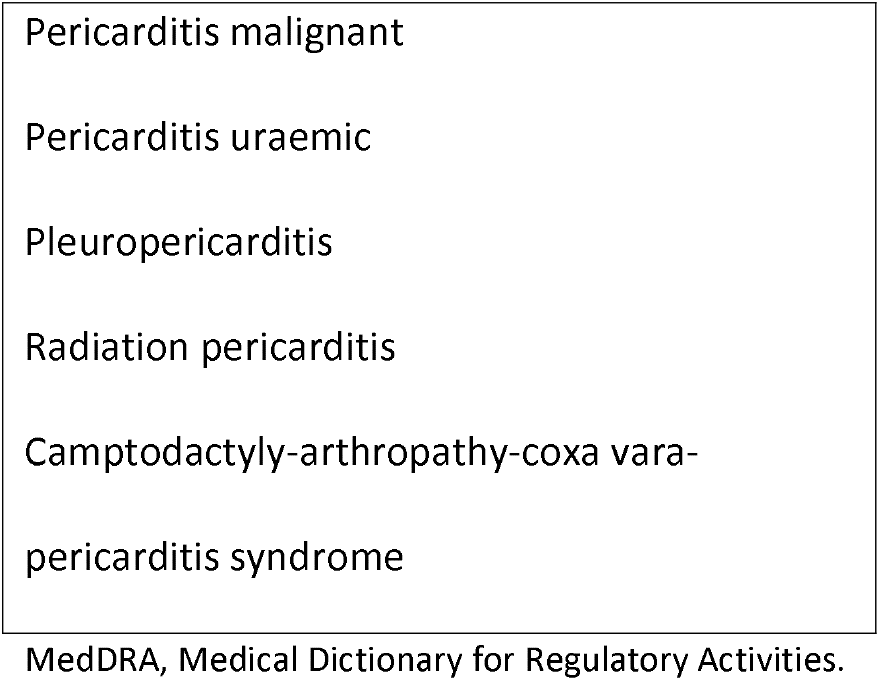
MedDRA Preferred Terms for Data Extraction.

## Notes

### Author Declarations

This study was based on data collected in the Moderna global safety database, mostly from adverse event reports submitted voluntarily, as part of routine ongoing post-authorization safety surveillance efforts by Moderna, Inc., as required by regulatory authorities; accordingly, a central institutional review board (IRB; Advarra) confirmed this study met criteria for an exemption from IRB oversight under 45 CFR 46.104(d)(4).

## REFERENCES

1. Baden LR, El Sahly HM, Essink B, et al. Efficacy and Safety of the mRNA-1273 SARS-CoV-2 Vaccine. N Engl J Med. Feb 4 2021;384(5):403–416. doi:10.1056/NEJMoa2035389

2. Polack FP, Thomas SJ, Kitchin N, et al. Safety and Efficacy of the BNT162b2 mRNA Covid-19 Vaccine. N Engl J Med. Dec 31 2020;383(27):2603–2615. doi:10.1056/NEJMoa2034577

3. Oliver SE, Gargano JW, Marin M, et al. The Advisory Committee on Immunization Practices’ Interim Recommendation for Use of Pfizer-BioNTech COVID-19 Vaccine - United States, December 2020. MMWR Morb Mortal Wkly Rep. Dec 18 2020;69(50):1922–1924. doi:10.15585/mmwr.mm6950e2

4. Oliver SE, Gargano JW, Marin M, et al. The Advisory Committee on Immunization Practices’ Interim Recommendation for Use of Moderna COVID-19 Vaccine - United States, December 2020. MMWR Morb Mortal Wkly Rep. Jan 1 2021;69(5152):1653–1656. doi:10.15585/mmwr.mm695152e1

5. Wu Q, Dudley MZ, Chen X, et al. Evaluation of the safety profile of COVID-19 vaccines: a rapid review. BMC Med. Jul 28 2021;19(1):173. doi:10.1186/s12916-021-02059-5

6. Montgomery J, Ryan M, Engler R, et al. Myocarditis Following Immunization With mRNA COVID-19 Vaccines in Members of the US Military. JAMA Cardiol. Oct 1 2021;6(10):1202–1206. doi:10.1001/jamacardio.2021.2833

7. Israeli Ministry of Health. Surveillance of Myocarditis (Inflammation of the Heart Muscle) Cases Between December 2020 and May 2021 (Including). June 2, 2021. Accessed October 18, 2021. https://www.gov.il/en/departments/news/01062021-03

8. World Health Organization. COVID-19 subcommittee of the WHO Global Advisory Committee on Vaccine Safety (GACVS) reviews cases of mild myocarditis reported with COVID-19 mRNA vaccines. May 26, 2021. Accessed October 18, 2021. https://www.who.int/news/item/26-05-2021-gacvs-myocarditis-reported-with-covid-19-mrna-vaccines

9. Centers for Disease Control and Prevention. COVID-19 VaST Work Group Report – May 17, 2021. May 17, 2021. Accessed October 18, 2021. https://www.cdc.gov/vaccines/acip/work-groups-vast/report-2021-05-17.html

10. Vogel GC-F J.;. Israel reports link between rare cases of heart inflammation and COVID-19 vaccination in young men. Science. June 1, 2021. Accessed October 18, 2021. https://www.science.org/content/article/israel-reports-link-between-rare-cases-heart-inflammation-and-covid-19-vaccination

11. Ali K, Berman G, Zhou H, et al. Evaluation of mRNA-1273 SARS-CoV-2 Vaccine in Adolescents. N Engl J Med. Aug 11 2021;doi:10.1056/NEJMoa2109522

12. Frenck RW, Jr., Dormitzer PR, Gurtman A. BNT162b2 Covid-19 Vaccine in Adolescents. Reply. N Engl J Med. Sep 30 2021;385(14):1343. doi:10.1056/NEJMc2113394

13. Klein NP, Lewis N, Goddard K, et al. Surveillance for Adverse Events After COVID-19 mRNA Vaccination. JAMA. Oct 12 2021;326(14):1390–1399. doi:10.1001/jama.2021.15072

14. Simone A, Herald J, Chen A, et al. Acute Myocarditis Following COVID-19 mRNA Vaccination in Adults Aged 18 Years or Older. JAMA Intern Med. Oct 4 2021;doi:10.1001/jamainternmed.2021.5511

15. Gargano JW, Wallace M, Hadler SC, et al. Use of mRNA COVID-19 Vaccine After Reports of Myocarditis Among Vaccine Recipients: Update from the Advisory Committee on Immunization Practices - United States, June 2021. MMWR Morb Mortal Wkly Rep. Jul 9 2021;70(27):977–982. doi:10.15585/mmwr.mm7027e2

16. Mevorach D, Anis E, Cedar N, et al. Myocarditis after BNT162b2 mRNA Vaccine against Covid-19 in Israel. N Engl J Med. Oct 6 2021;doi:10.1056/NEJMoa2109730

17. Witberg G, Barda N, Hoss S, et al. Myocarditis after Covid-19 Vaccination in a Large Health Care Organization. N Engl J Med. Oct 6 2021;doi:10.1056/NEJMoa2110737

18. Brighton Collaboration. Myocarditis/pericarditis case definition. The Task Force for Global Health, July 16, 2021. https://brightoncollaboration.us/myocarditis-case-definition-update/

19. World Health Organization. Causality assessment of an Adverse Event Following Immunization (AEFI): User Manual for the Revised WHO Classification. Accessed October 18, 2021. https://www.who.int/vaccine_safety/publications/aevi_manual.pdf?ua51

20. Centers for Disease Control and Prevention. COVID-19 Vaccinations in the United States. Accessed October 1, 2021. https://covid.cdc.gov/covid-data-tracker/#vaccinations_vacc-total-admin-rate-total

21. European Centre for Disease Prevention and Control. European Centre for Disease Prevention and Control COVID-19 Vaccine Tracker. Accessed October 1, 2021. https://vaccinetracker.ecdc.europa.eu/public/extensions/COVID-19/vaccine-tracker.html

22. Public Health Agency of Canada. COVID-19 vaccination in Canada. Accessed October 1, 2021. https://health-infobase.canada.ca/covid-19/vaccination-coverage/

23. Federal Office of Public Health Switzerland. COVID-19 Switzerland. Accessed October 1, 2021. https://www.covid19.admin.ch/en/epidemiologic/vacc-doses

24. Ritchie H, Mathieu E, Rodes-Guirao L, et al. Coronavirus Pandemic (COVID-19). Published online at http://OurWorldInData.org. Accessed October 1, 2021. https://ourworldindata.org/coronavirus

25. Gubernot D, Jazwa A, Niu M, et al. U.S. Population-Based background incidence rates of medical conditions for use in safety assessment of COVID-19 vaccines. Vaccine. Jun 23 2021;39(28):3666–3677. doi:10.1016/j.vaccine.2021.05.016

26. Fairweather D, Cooper LT, Jr., Blauwet LA. Sex and gender differences in myocarditis and dilated cardiomyopathy. Curr Probl Cardiol. Jan 2013;38(1):7–46. doi:10.1016/j.cpcardiol.2012.07.003

27. Shimabukuro T. COVID-19 Vaccine Safety Updates: Advisory Committee on Immunization Practices (ACIP). June 23, 2021. Accessed October 1, 2021. https://www.cdc.gov/vaccines/acip/meetings/downloads/slides-2021-06/03-COVID-Shimabukuro-508.pdf

28. Barda N, Dagan N, Ben-Shlomo Y, et al. Safety of the BNT162b2 mRNA Covid-19 Vaccine in a Nationwide Setting. N Engl J Med. Sep 16 2021;385(12):1078–1090. doi:10.1056/NEJMoa2110475

29. Su JR, McNeil MM, Welsh KJ, et al. Myopericarditis after vaccination, Vaccine Adverse Event Reporting System (VAERS), 1990-2018. Vaccine. Jan 29 2021;39(5):839–845. doi:10.1016/j.vaccine.2020.12.046

30. Bozkurt B, Kamat I, Hotez PJ. Myocarditis With COVID-19 mRNA Vaccines. Circulation. Aug 10 2021;144(6):471–484. doi:10.1161/CIRCULATIONAHA.121.056135

31. Halsell JS, Riddle JR, Atwood JE, et al. Myopericarditis following smallpox vaccination among vaccinia-naive US military personnel. JAMA. Jun 25 2003;289(24):3283–9. doi:10.1001/jama.289.24.3283

32. Pollack A, Kontorovich AR, Fuster V, Dec GW. Viral myocarditis--diagnosis, treatment options, and current controversies. Nat Rev Cardiol. Nov 2015;12(11):670–80. doi:10.1038/nrcardio.2015.108

33. Boehmer TK, Kompaniyets L, Lavery AM, et al. Association Between COVID-19 and Myocarditis Using Hospital-Based Administrative Data - United States, March 2020-January 2021. MMWR Morb Mortal Wkly Rep. Sep 3 2021;70(35):1228–1232. doi:10.15585/mmwr.mm7035e5

34. Singer ME, Taub IB, Kaelber DC. Risk of Myocarditis from COVID-19 Infection in People Under Age 20: A Population-Based Analysis. medRxiv. Jul 27 2021;doi:10.1101/2021.07.23.21260998

35. Wong H-L. Surveillance Updates of Myocarditis/Pericarditis and mRNA COVID-19 Vaccination in the FDA BEST System. October 14, 2021. Accessed October 15, 2021. https://www.fda.gov/media/153090/download

36. Talbot HK, Hopkins RH. COVID-19 Vaccine Safety Technical (VaST) Work Group: Safety Assessments. October 21, 2021. Accessed October 21, 2021. https://www.cdc.gov/vaccines/acip/meetings/downloads/slides-2021-10-20-21/09-COVID-Talbot-508.pdf

37. World Health Organization. COVID-19 subcommittee of the WHO Global Advisory Committee on Vaccine Safety (GACVS): updated statement regarding myocarditis and pericarditis reported with COVID-19 mRNA vaccines. October 27, 2021. Accessed October 28, 2021. https://www.who.int/news/item/27-10-2021-gacvs-statement-myocarditis-pericarditis-covid-19-mrna-vaccines-updated

38. Su J. Myopericarditis following COVID-19 vaccination: Updates from the Vaccine Adverse Event Reporting System (VAERS). October 21, 2021. Accessed October 21, 2021. https://www.cdc.gov/vaccines/acip/meetings/downloads/slides-2021-10-20-21/07-COVID-Su-508.pdf

39. Klein N. Myocarditis Analyses in the Vaccine Safety Datalink: Rapid Cycle Analyses and “Head-to-Head” Product Comparisons. Accessed October 21,, 2021. https://www.cdc.gov/vaccines/acip/meetings/downloads/slides-2021-10-20-21/08-COVID-Klein-508.pdf

40. Tang P, Hasan MR, Chemaitelly H, et al. BNT162b2 and mRNA-1273 COVID-19 vaccine effectiveness against the Delta (B.1.617.2) variant in Qatar. MedRxiv. 2021;2021.08.11.21261885 doi:doi: https://doi.org/10.1101/2021.08.11.21261885

41. Gram MA, Nielsen J, Schelde AB, et al. Vaccine effectiveness when combining the ChAdOx1 vaccine as the first dose with an mRNA COVID-19 vaccine as the second dose. MedRxiv. 2021;2021.07.26.21261130 doi:https://doi.org/10.1101/2021.07.26.21261130

42. Puranik A, Lenehan PJ, Silvert E, et al. Comparison of two highly-effective mRNA vaccines for COVID-19 during periods of Alpha and Delta variant prevalence. medRxiv. Aug 9 2021;doi:10.1101/2021.08.06.21261707

43. Nordstrom P, Ballin M, Nordstrom A. Effectiveness of heterologous ChAdOx1 nCoV-19 and mRNA prime-boost vaccination against symptomatic Covid-19 infection in Sweden: A nationwide cohort study. The Lancet Regional Health-Europe. 2021;doi:https://doi.org/10.1016/j.lanepe.2021.100249

44. Das BB, Kohli U, Ramachandran P, et al. Myopericarditis after messenger RNA Coronavirus Disease 2019 Vaccination in Adolescents 12 to 18 Years of Age. J Pediatr. Jul 30 2021;doi:10.1016/j.jpeds.2021.07.044

